# Social-distancing effectiveness tracking of the COVID-19 hotspot Stockholm

**DOI:** 10.1101/2020.06.30.20143487

**Authors:** Joachim Oberhammer

**Affiliations:** Department of Intelligent Systems, KTH Royal Institute of Technology, Malvinas väg 10, SE-100 44 Stockholm, Sweden

## Abstract

**Background:** The COVID-19 outbreak in Stockholm, Sweden, is characterized by a near-absence of governmental interventions and high fatalities in the care home population. This study analyses the outbreak and the social-distancing effectiveness timeline in the general population and the care homes.

**Methods:** A novel distributed-compartmental, time-variant epidemiological model was designed specifically for COVID-19 transmission characteristics, featuring a/pre/symptomatic transmission, a non-linear hospital model, a weakly-coupled sub-model for the care-home population, and parametrized continuous social-distancing functions. The model parameters and the social-distancing timelines are determined by randomization and Monte-Carlo simulations analysing real-world data.

**Findings:** Despite a high initial reproduction number (3·29) and the near-absence of governmental interventions, the model quantitated that the transmission rate in the general population was suppressed by 73%, and in the care homes by 79%. The measures in the care homes took effect 4·8 days delayed; and if applied 4 or 8 days earlier, the fatalities could have been reduced by 63·2% or 89·9%. The infected population is estimated to 16·2% (June 10). An expected underestimation of population immunity by antibody studies is confirmed. The infection fatality ratio extrapolates to 0·61% (peak: 1·34%). The model indicates a seasonal effect which effectively suppressed a new rise. An analysed large-scale public event had no large influence. The asymptomatic ratio was determined to 35%.

**Interpretation:** The proposed model and methods have proven to analyse a COVID-19 outbreak and to re-construct the social-distancing behaviour with unprecedented accuracy, confirming even minor details indicated by mobility-data analysis, and are applicable to other regions and other emerging infectious diseases of similar transmission characteristics. The self-regulation of the population in Stockholm, influenced by advices by the authorities, was able to suppress a COVID-19 outbreak to a level far beyond that the stringency index of governmental interventions suggests. Proper timing of effective measures in the care homes is important to reduce fatalities.

## Introduction

Sweden, has never imposed a lockdown and only issued very few governmental restrictions. Instead, the population has been briefed daily with advices by the Public Health Agency of Sweden (Folkhälsomyndigheten, FHM). As of June 10, 4795 confirmed COVID-19 deaths have been reported in Sweden (population: 10·3 million), by that time the fifth highest death rate per capita in the world. Sweden has a 80·3% share of the deaths in North Europe, despite only having 38·2% of the Nordic population.^1^ The region Stockholm (23% of the population of Sweden) accounts for 45·4% of all deaths in Sweden, and is characterized by a large proportion of deaths (43·3%) having occurred in the care-homes population (only 0·6% of the total population).

Epidemiological modelling of the COVID-19 outbreak has been used for forecasting the healthcare demand^2^ and to develop containment strategies using non-pharmaceutical interventions (NPI).^3,4,5^ Modelling is also used to determine key parameters such as population immunity and infection fatality rate (IFR), and to analyse the impact of imposing and revoking social-distancing measures.^6,7,8,9,10^ A well-established class of models is the Susceptible(S)-Exposed(E)-Infected(I)-Recovered(R)-Deceased(D) compartmental model.^11,12,13^ Due to short simulation times, in particular deterministic SEIRD models are used for parameter estimation by randomization.^14^ The impact of interventions is usually modelled by discrete steps of the infectivity parameter.^2,6,8,15^ It was found that multiple inferring change points are more suitable for tracking the effect of specific interventions.^7^ A continuous modelling of the transmission-rate reduction is assumed to be more suitable for countries like Sweden, without disruptive governmental interventions and adjusting to the self-regulation behaviour of the population.

## Methods

### Epidemic simulation model

The deterministic, time-variant SEIRD model (Figure 1) features distributed compartments for more realistic modelling of the different phases of the infection by time-distributed parameters (infectivity, hospitalization and death rates, probabilities for PCR or antibody positive testing) as shown in Figure 2:A. The model comprises two weakly coupled sub-models, for modelling the different dynamics of the general population (GP) and the care homes (CH). The GP model features an asymptomatic (individuals never developing clearly noticeable symptoms) and a symptomatic branch, with the latter structured into pre-symptomatic and symptomatic compartments. Only individuals in compartments I_1_—I_4_ are considered infectious in both branches, and only compartments I_3_-I_6_ of the symptomatic branch have associated hospitalization and death rates. Environmental transmission is not modelled due to its minor contribution for COVID-19.^16,17,18^ Age-groups, work/household environment, or individual susceptibility is not modelled.^19,20^

**Figure 1.**
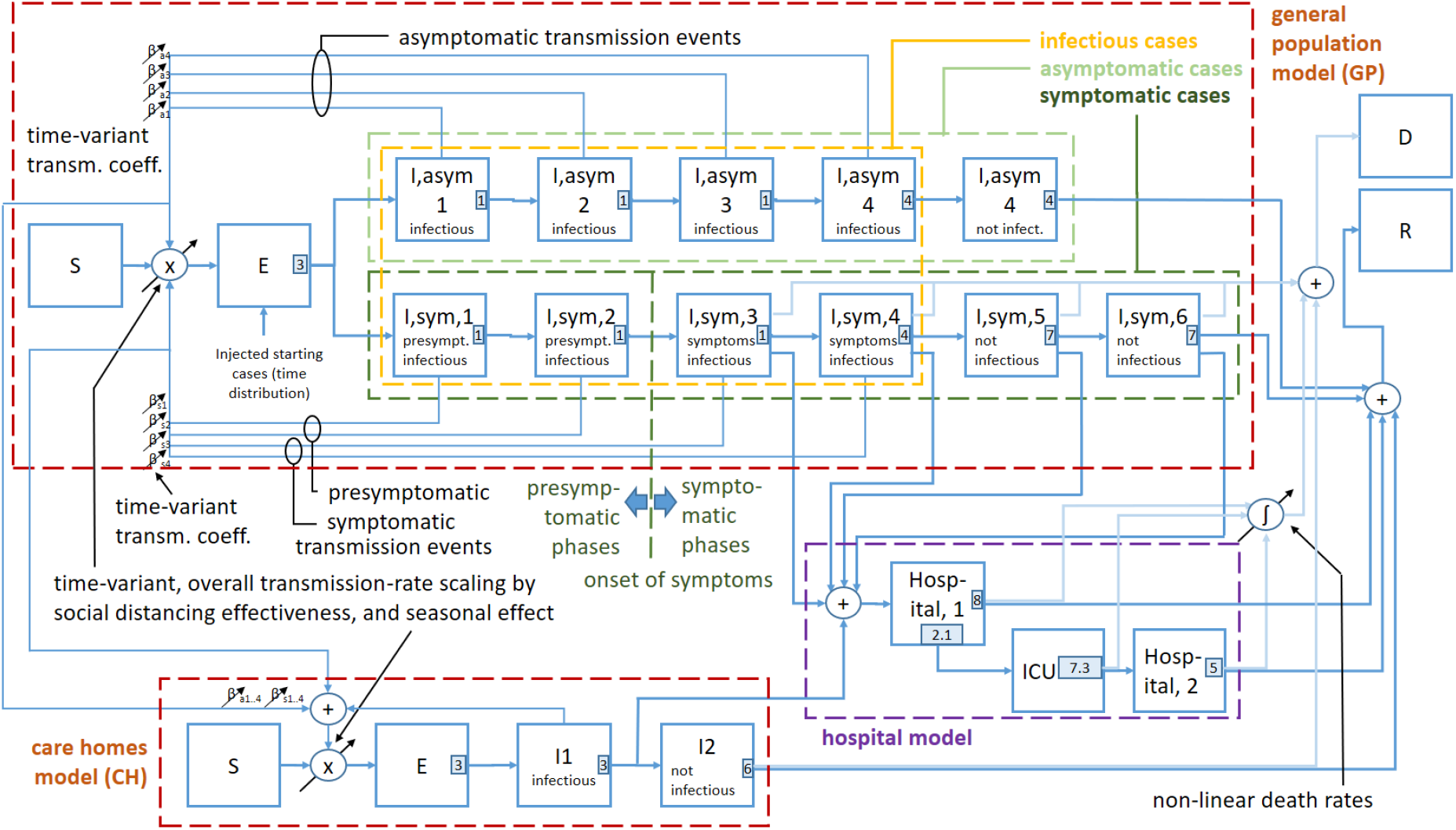
Flow chart of the deterministic, time-variant, distributed-compartmental model with a detailed sub-model for the general population including a symptomatic and an asymptomatic branch, a weakly-coupled sub-model for the care-home population, and a common hospital model.

**Figure 2.**
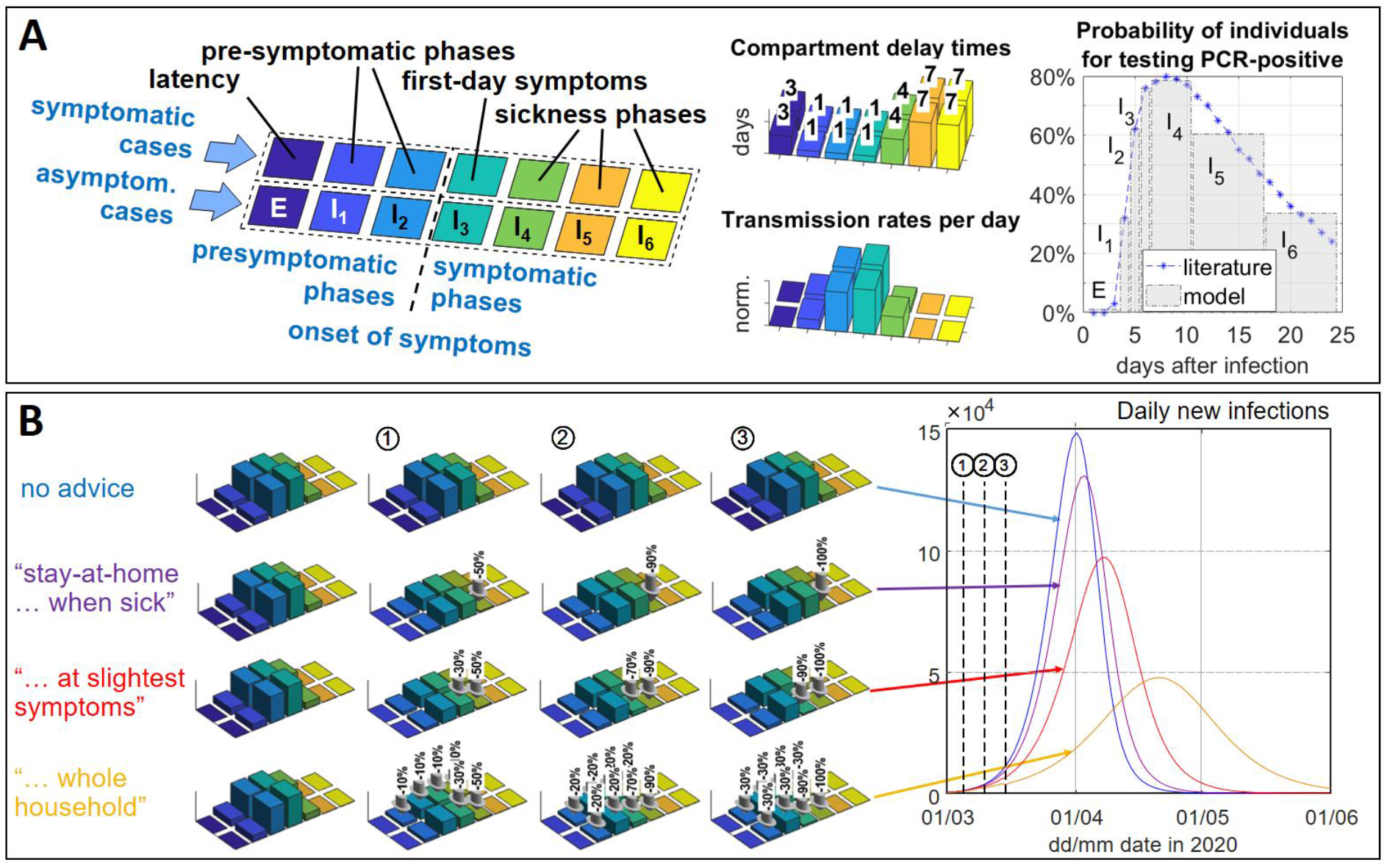
Compartments with time-distributed infection parameters of the general population model: A: asymptomatic and symptomatic branches with associated key parameters (delay times; transmission rates; probability for positive RT PCR tests based on Mizumoto et al.^30^). B: Modelling of the effect of advices by Swedish authorities, by reducing the infectivity of different compartments (the last advice, whole household quarantine, was never issued in Sweden).

For the GP model, the force of infection *λ*_*GP*_ (in Δ*S*_*GP*_ = −*λ*_*GP*_·*S*_*GP*_) is as follows:

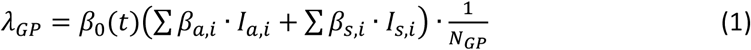

(*S*_*GP*_ susceptible population; *I*_*a,i*_ and *I*_*s,i*_ individuals and *β*_*a,i*_ and *β*_*s,i*_ transmission rates in asymptomatic/symptomatic branch compartments; *β*_0_(*t*) time-variant transmission-rate matrix scaling factor: ***β***_*GP*_ = *β*_0_(*t*) [*β*_*a,i*_; *β*_*s,i*_]; *N*_*GP*_ host population, Stockholm: 2374550).

The CH model is simplified (3-day latency; 3-day infectious phase I_1_ including two presymptomatic and a symptomatic day; 7-day sickness phase I_2_ without transmission, i.e. patient isolation assumed), without distinguishing asymptomatic/symptomatic branches, since CH population is negligible (0·6% of GP) for a transmission event analysis. Besides the internal factor *α*_*INT*_, the force of infection *λ*_*CH*_ s weakly coupled to the GP model by an external factor *α*_*EXT*_ modelling imported transmissions by health care workers and visitors:

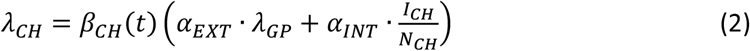

(*I*_*CH*_ infectious individuals in I_1_; *N*_*CH*_ CH population size, 15000 estimated,^21^ time-variant factor *β*_*CH*_(*t*)). Transmission-events feedback from the CH to the GP model is assumed neligible.

The hospital model comprises a primary ward, an intensive care, and a secondary general-ward compartment for severely-sick, critically-sick, and patients recovering from intensive care, respectively.

### Time-variant and non-linear model parameters

The GP and CH transmission rates are scaled by the time-variant factors *β*_0_ and *β*_*CH*_, in the following referred to as social-distancing effectiveness functions (SDEF), which track the social-distancing behaviour and seasonal effects. All elements of the transmission-rate matrix are independently controllable by time-dependent reduction factors, which allows to investigate the effect of different advices to the population, from staying-home with symptoms to whole-household quarantine (Figure 2:B).

Since the reported number of patients in ICU in Stockholm follows not exactly the shape of the general ward, it was assumed that with time a growing proportion of patients can be kept in regular ward, which was implemented by a three-parameter logistics function (details in Appendix). The hospital death rate, in relation to the hospital occupancy, was found to decrease with time, which might reflect increasing treatment experience or a reduced number of CH patients transferred to hospital, which is also modelled by a parametrized logistics function. A non-linear hospital and ICU death-rate scaling factor, depending on the hospital occupancy, is also implemented by a logistics function.

### SARS-CoV-2 infection parameters

The onset of symptoms after infection is set to 5 days (discretization of the 5·2 days incubation time).^22,23^ The distribution of the transmission rates in the two presymptomatic (I_1_, I_2_) and the two symptomatic (I_3_, I_4_) infectious compartments (Figure 2:A) is following the distribution of transmission-pair analysis so that the proportion of the presymptomatic transmission events in the symptomatic branch is 44%.^24^ The total contagious period (I_1_-I_4_) is 7 day.^22^ Since literature reports no significant difference in viral spread between asymptomatic and symptomatic individuals,^25,26^ the initial infectivity in the two branches is the same, followed by a 75% reduction after the first symptomatic day in the symptomatic branch when symptomatic individuals are assumed to reduce social interaction.

The proportion of cases moving into the asymptomatic branch is determined by analysing the proportion of non-symptomatic (asymptomatic and presymptomatic) individuals testing PCR-positive, taking the dynamics of the outbreak into account, and comparing to an average reported in the literature (pre- and post-lockdown study in Vo, Italy;^25^ Diamond Princess;^27^ Heinsberg study;^28^ repatriating individuals to the UK;^2^ average: 41·5%; Figure 3). The best matching asymptomatic-branch proportion is 35%, assuming that *R*_*eff*_ in the studies is around 1·0.

**Figure 3.**
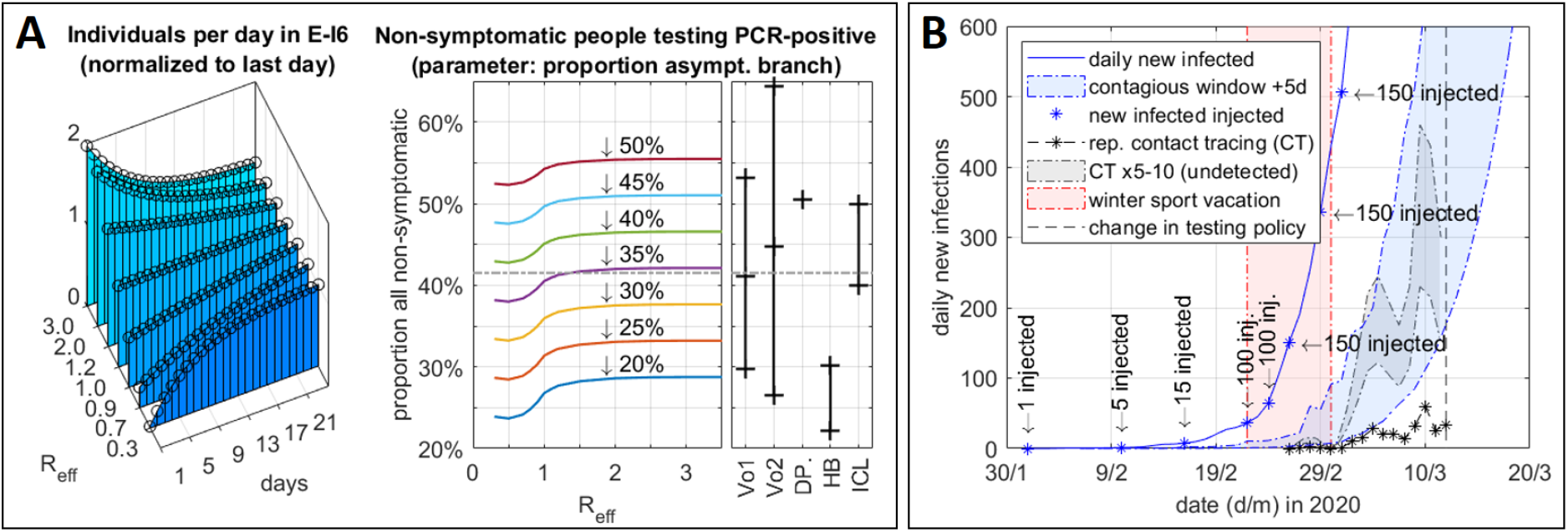
A: Estimation of the asymptomatic branch ratio, by analysing the proportion of identified non-symptomatic individuals to the literature (Vo,1 and Vo,2: pre- and post-lockdown study in the Italian city of Vo;^25^ DP: study from cruising ship *Diamond Princess*;^27^ Heinsberg study, range as given in the study for no symptoms and 1 out of 15 symptoms;^28^ ICL: repatriating individuals to the UK^2^). B: Starting sequence of the model, mapping the reported contact-tracing cases, multiplied by a dark-figure factor of 5—10, and assuming a 5-day reporting delay, to the daily new infections in the model.

Antibody development in the model is assumed to happens within 1 to 3 weeks after infection^29^ using a normal distribution (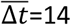 days, 95%CI[4·5, 23·5]). The mapped probability distribution of individuals to test PCR positive in different phases^30^ is shown in Figure 2:A. For the care-home model (CH), the outcome is decided in average seven days after onset of symptoms (FHM: two thirds of all CH fatalities in the first week). The CH death rate is 39·75% (reported CFR for CH, Stockholm, May 25). The hospital transfer rate is set to 5% (assumption based on: only 10·7% of all care-home inhabitants in Sweden are younger than 75 years;^31^ and FHM: only a small fraction of care-home patients benefits from critical and intensive care), which is also reflected by the high proportion of CH deaths and the low ICU death rate.

The ICU is modelled after a report by the Swedish Intensive-Care Register (SIR) of April 30 (median/mean/max time: 6·75/7·25/31·8 days; 29·75% death rate; time on ward before transfer to ICU: 2·1 days). ^32^ The average times on regular (H_1_) and on post-ICU (H_2_) ward are 8 and 5 days, respectively.^33^ It is assumed that most people surviving ICU recover, therefore the H_2_ death rate is arbitrarily set to 5% of the H_1_ death rate. The hospitalization-rate distribution, i.e. the transfer from the GP symptomatic-branch compartments, was set so that the average time from onset of symptoms to hospitalization is 8·16 days.^32^

### Model fitting to real-world data: social-distancing tracking

The starting values and randomization confidence intervals of the unknown model parameters (SDEF for GP and CH; hospitalization rate; ward-to-ICU transfer, hospital death-rate, and the hospital non-linearity parameters) were determined by sensitivity analysis using multi-dimensional parameter variation, with the GP SDEF initially modelled by a three-parameter logistics function. The parameters were then refined by randomization with Monte-Carlo simulations. Minimum mean square error functions were used to benchmark the model outcome to real-world data: the number of patients in critical care and in intensive care, the number of date-adjusted GP and CH deaths (daily situation reports, Region Stockholm), and the results of two PCR studies by FHM in April.^34,35^ The results of four weekly antibody studies by FHM (April to June),^36^ where non-COVID-19 patients seeking ambulatory care in Stockholm were sampled, was not included for model fitting since FHM reported on May 26 a potential bias towards persons with a lower risk of infection for this type of sampling, and since in particular mild/asymptomatic people do not have a robust antibody response. ^26,37^

The model is started by injecting infected cases to the latency compartment so that the daily new infections are matching the reported confirmed cases from contact tracing, assuming a 5-day reporting delay and a 5—10 times larger dark number range (Figure 3:B). The injections happen primarily in Stockholm’s winter school vacation week, reported by FHM to have been the major source of imported cases.

For the determination of the SDEF function of the GP by Monte-Carlo simulations, 15 randomized points with shape-preserving piecewise cubic interpolation were mapped on the initial logistics function. The points were optimized by three subsequent Monte-Carlo batches with 1 million simulation runs each, where the 95%CI calculated from the corrected sample standard deviation of the 100 best functions of a previous batch were used as the 95%CI for the randomization in a subsequent batch (convergence shown in Figure 4:A). The optimization of the hospital parameters was done by a subsequent 1-million-run batch (Figure 4:C). For the CH model, the SDEF was modelled as a three-parameter logistic function, determined with the internal and external transmission-rate factors by a 1-million simulation-run batch. All parameters were fine-adjusted with a combined 1-million run batch. More details on the model and data fitting in the Appendix.

**Figure 4.**
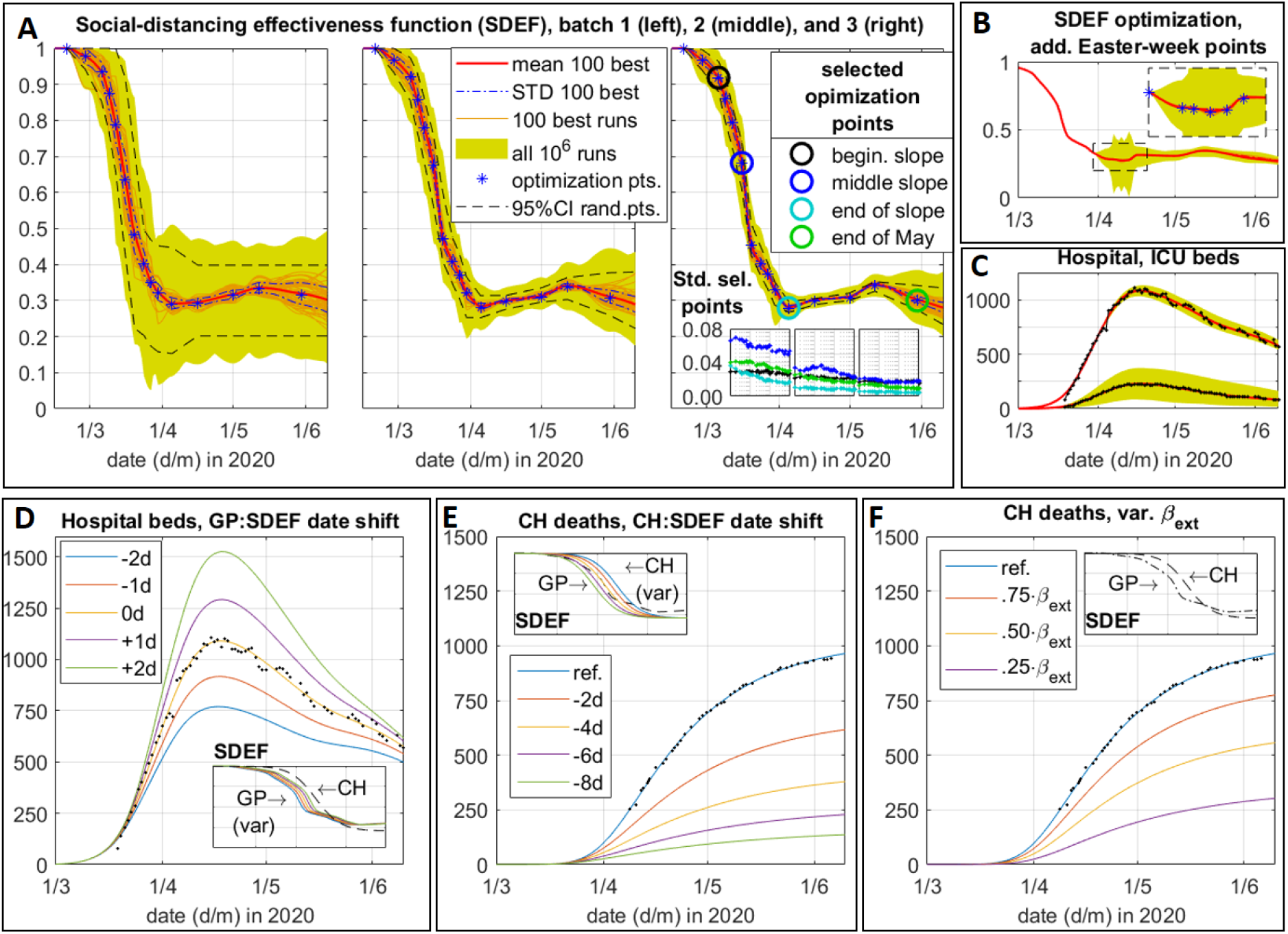
Real-world data fitting results by Monte-Carlo simulations (A-C), and outcome analysis by parameter variation (D-F). A: social-distancing effectiveness function (SDEF) determined by three simulation batches with 1 million runs each (convergence of standard-deviation of selected points shown in the insert); B: results of 2 million runs with additional optimization points for refining the Easter-week minimum; C: optimization of ICU transfer rate; D: effect of time shift of SDEF of GP by ±2 days; E: time shift of SDEF of CH; F: effect of reduction of the infections by externals into the CH. (GP=general population; CH=care homes).

## Results

### Mapping the outbreak in Stockholm

The SDEF determined for the GP with the advices to the population by FHM, governmental measures, and key events of the outbreak mapped on the timeline, with comparison to Google’s mobility data analysis^38^ and the Oxford Governmental Stringency Index,^15^ is shown in Figure 5. From an initial 1·0 (February 1), the function falls to a minimum of 0·29 (April 5). The major reduction happens between March 6 and 20, when the majority of the governmental restrictions and advices were issued. The slope and the increase after April 4 are in good agreement with the mobility data, which recorded for Stockholm a major slope between March 7 and March 19 (−31% retail/recreation mobility; -45% public-transport; -42% workplace-mobility) and shows a minimum for the Easter weekend, for which media reported a traffic reduction by 90% as compared to previous years. A more detailed mapping of the Easter week (April 5-15) was carried out by three additional randomization points, while randomizing also all surrounding and following points in two 1-million simulation-run batches (Figure 4:B), revealing a 3·9% improved error-function, a deeper SDEF minima to 0·27, and a rise thereafter to the previous level (comparison of the two SDEF in Figure 5). After a local peak (May 13; 0·34) the social distancing function falls again, contrary to the mobility data. A time-shifting of the SDEF by ±2 days results in a large variation in peak hospital beds (−763 (−29%) to 1517 (+39%); Figure 4:D), which confirms the previously reported sensitivity to the timing of NPIs.^7^ Verification tests of the SDEF and its independency on the starting point choice were done by shifting the starting points ±1—±3 days (500000 Monte-Carlo runs each). The SDEF function was confirmed by all verification functions, with the local minima (April 5) and the local maxima (May 13) of the original SDEF matching the 95%CI (estimated from the 100 best functions each) of 5 out of 6 verification functions. The average deviation of the 50% transition was 0·32 days (95%CI[0·28, 0·36]) (details in Appendix).

**Figure 5.**
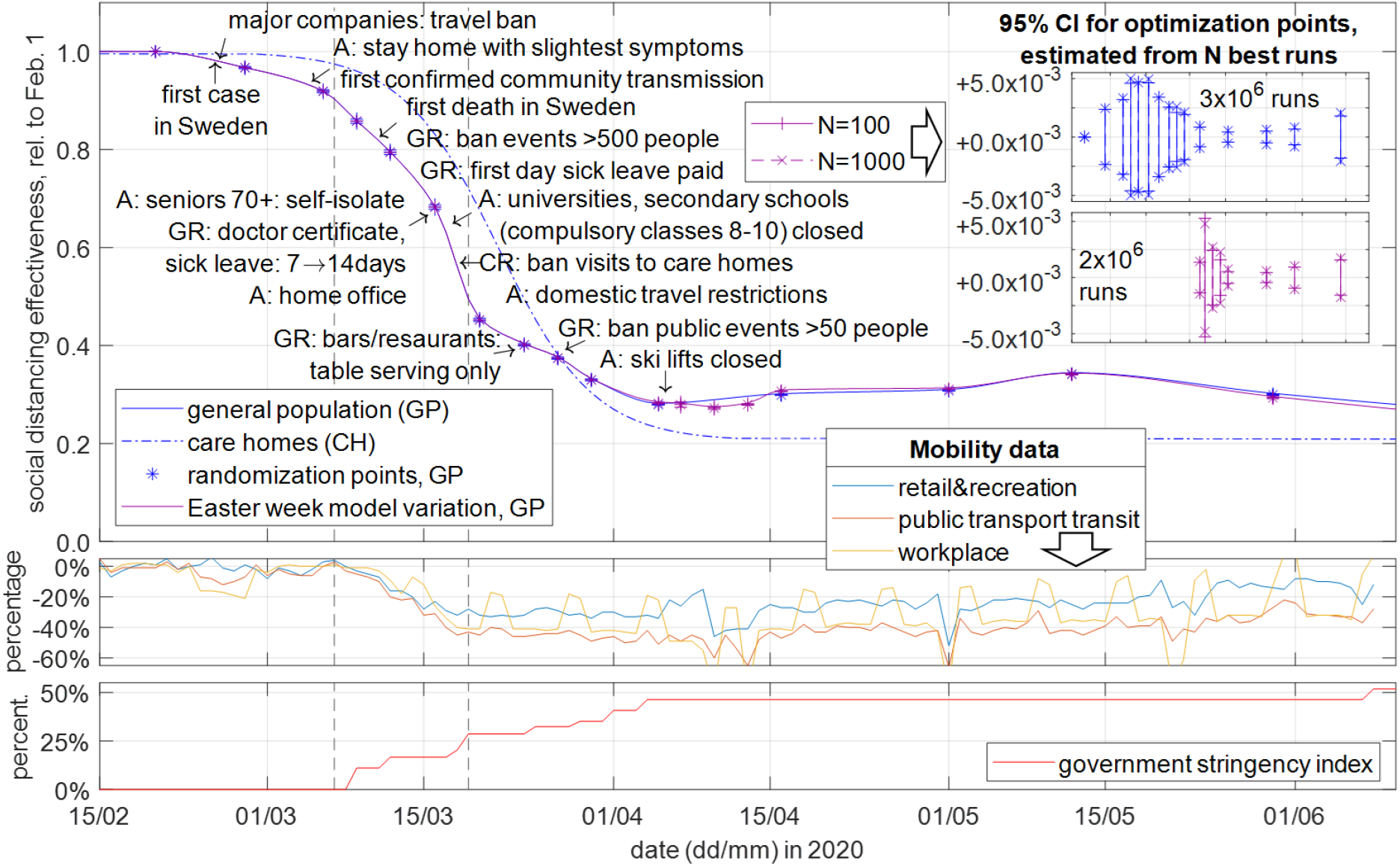
Social distancing effectiveness function (SDEF) determined by the proposed model and Monte-Carlo simulations data-fitting, with advices and governmental restrictions mapped on the timeline (A=advice by Public Health Agency; GR=governmental restriction; CR=care-home restriction), including a SDEF variation with additional randomization points for resolving the Easter week minima indicated by the mobility data; middle: Google mobility data for Stockholm;^38^ bottom: Oxford government stringency index for Sweden.^15^

The delay between the SDEFs of the CH to the GP was determined to 4·8 days (95%CI[4·67, 4·92]), and the final level is lower for the CH (0·2048, 95%CI[0·2038, 0·2058]) than for the GP, indicating that measures in the CH were taken later but were more effective. Figure 4:E shows that the CH deaths could have been reduced by 63·2% (606 people, 95%CI[604, 608]) if the transmission-rate reduction in the CHs would have been simultaneously with the GP, or, if additional four days earlier, even by 89·9% (95%CI[89·6, 90·2]). If the imported infections by CH workers and visitors would have been halved early on, the CH deaths could have been reduced by 44·6% (95%CI[44·4, 44·6]; Figure 4:F). With 43·3% of Stockholm’s total deaths attributed to the care homes, these reductions would have decreased the total fatalities by 15·9%, 43·7%, and 24%, respectively. The hospital death-rate nonlinearity for peak day (April 6) was determined to +6·6% (95%CI[-5·5, 18·8]). By June 10, 16·2% (95%CI[15·9, 16·48]) of the hospital patients had needed intensive care.

Figure 6 summarizes the model results, showing an excellent fit to the real-world data (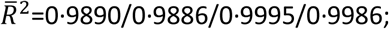 internally-studentized residuals shown). The basic reproduction number *R*_*0*_ was determined to 3·29, by replacing the starting sequence with an equivalent single injection (Figure 6:F). *R*_*eff*_, after falling below 1·0 on March 27 and reaching a minimum of 0·84 on April 1, increases to 0·99 on May 7, after which it falls again. The confidence intervals given in the following, for data referring to the total number of infected, are dominated not by the data-fitting but by the PCR-study reference-points uncertainty. By June 10, 16·2% (95%CI[10·9, 22·8]) of the population in Stockholm had been infected, increasing by 0·8% (95%CI[0·54, 1·12]) per day (Figure 6:D). The population immunity predicted by the model matches the results of FHM’s antibody studies at the upper quartile of their confidence-intervals (Figure 6:D), confirming the expected underestimation (average difference: -1·57%, 95%CI[-1·06, -2·22]). The number of infectious people has been reduced from its peak of 44140 (95%CI[29680, 62269]; April 1) to 15120 (95%CI[10167, 21330]), and the daily new infections are down from a peak of 6719 (95%CI=[4518, 9478), March 23) to 1590 (95%CI[1069, 2243]), same level as of March 8 (Figure 6:E). The hospitalization rate was 2·27% (95%CI[1·52, 3·20]). The peak infection fatality rate (IFR) was 1·34% (95%CI[0·90, 1·89]), and would extrapolate to 0·61% (95%CI[0·41, 0·86]; Figure 6:F). The asymptomatic ratio is determined to 35% by analysing the literature with this model.

**Figure 6.**
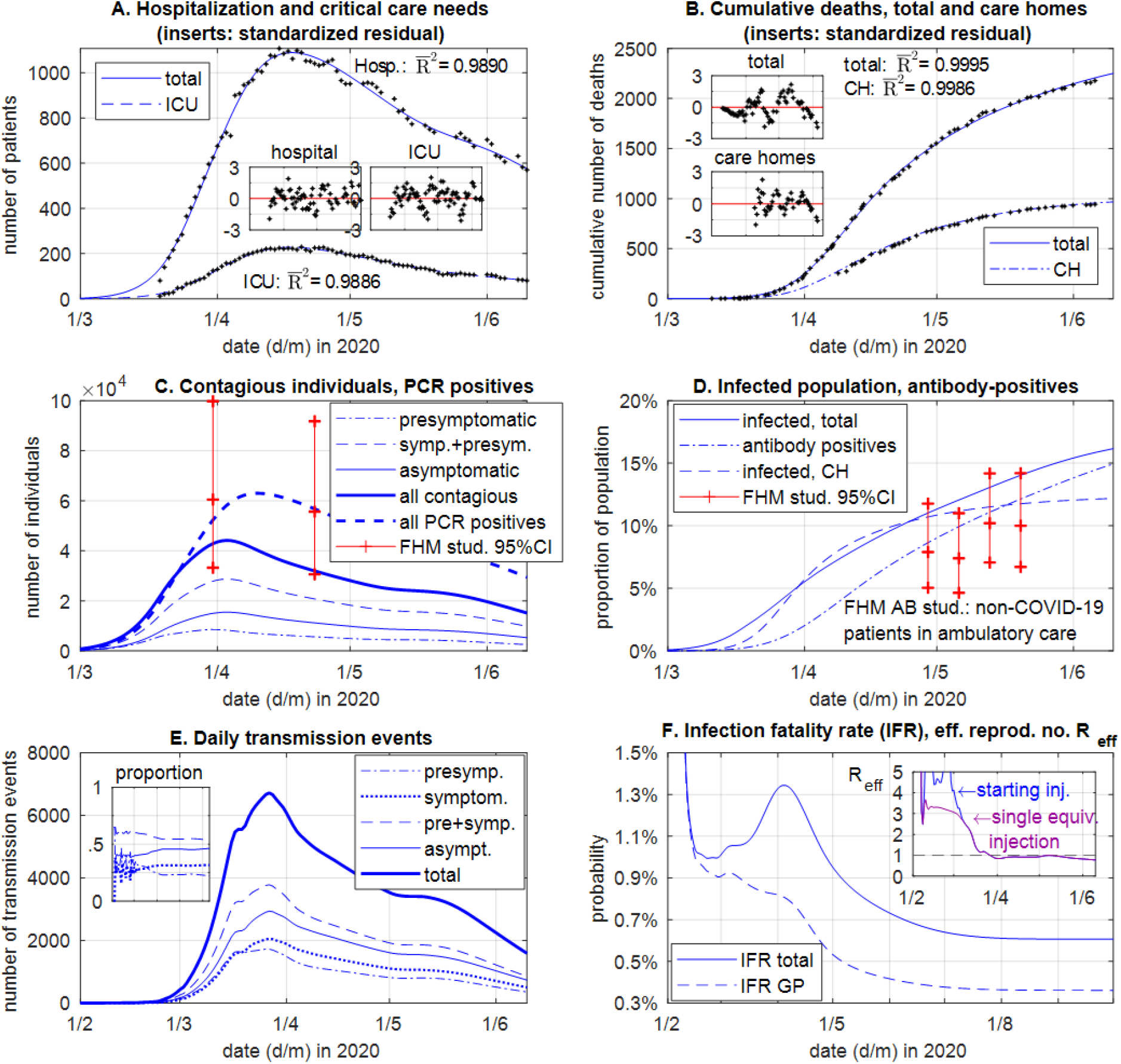
Simulation results of the model mapped to the real-world data of the COVID-19 outbreak in Stockholm until June 10, 2020. (GP=general population, CH=care home population)

### Europe’s last pre-corona large-scale public event: the Eurovision Song Contest final in Stockholm

The Eurovision Song Contest (ESC) Swedish Final (Stockholm, March 7, 27000 attendees), took place at a time when Sweden was the only country in Europe which had not banned large-scale public gatherings. Several COVID-19 super-spreader are known,^28,^39 and can have severe impact an outbreak, as in South Korea where 61·3% of all cases by March 15 were associated with a church service on February 9.^40^ The model determines the epidemiological parameters in Stockholm on March 7 to: *R*_*eff*_=2·76, 6674 infected, 2011 contagious, 1449 daily new infections, 23·3 contagious people present at the event. The potential impact of the event was analysed by injecting an average number *N* of transmissions per contagious person, scaling the model starting sequence to maintain same outcome, and then removing the injected ESC incidences to determine the outcome without the event. For *N*=5/10/20, the ESC contributes by 115/226/439 additional transmissions, and, if cancelled or conducted without public, the peak hospital occupancy would have been reduced from 1084 to 1069/1055/1028 patients (−1·4/-2·7/-5·4%). A local raise in the number of hospital patients, even for *N*=50 determined to only 4·6 patients, is not visible in the real-world data with a localized (17 points) standard-deviation of 11·4 patients. If happened on February 2 or 3 weeks earlier, and a single contagious individual present would have triggered 20 or 50 transmission-chains, the event would have added 12·9% or 30·8% to the epidemic, respectively.

## Discussion

The proposed distributed-compartmental epidemiological model, with COVID-19 specific infection characteristics including a sub-model for the care homes, was able to track the Stockholm outbreak and to quantitate the social-distancing effectiveness function with an unprecedented accuracy so that even small details in the timeline, indicated by mobility data analysis, were resolved. The model is constructed as a trade-off between model complexity, accuracy, and speed for suitability for large-scale data fitting methods. This study has quantitated that by primarily relying on the advices by authorities and on the self-regulation of the population, as in Sweden, it is possible to at least temporarily suppress COVID-19 by 73%, far beyond the government stringency-index indication. The determined social-distancing behaviour of the general population follows the mobility data analysis, whereas the determined care homes transmission-rate reduction, delayed by several days, follows the delayed government stringency index timeline. This indicates that the general-population self-regulation effect was ahead of time of the effect of governmental measures. The decrease of the social-distancing function after a local peak in May, contradicted by mobility data, indicates the onset of a summer seasonal effect and has prevented *R*_*eff*_ from climbing again above 1·0. The model outcome, in agreement with previous studies, is very sensitive to the timing of the social-distancing function. An anticipated underestimation of the population immunity by antibody studies is confirmed. This study has quantitated the importance of applying early care-home protection measures. The analysis of Europe’s last large-scale public event in pre-corona times has found that at an already wide-spread community transmission such an event has a significantly lower impact as compared to happening in a more sensitive earlier phase. The proposed model and methods are applicable to other regions and other emerging infectious diseases with similar transmission characteristics as COVID-19.

## Data Availability

The source code of this model is enclosed in the appendix. The source code and the database with model parameters will be made freely available on https://github.com/JoachimKTH/COVID19_SEIRD_model.

## Acknowledgements

Peter L E Konings, Jasmine Gardner, Nele Brusselaers, Wouter Metsola van der Wijngaart, are acknowledged for a first feedback on the simulation model.

